# Cerebellar anaplastic astrocytoma in adult patients: 15 consecutive cases from a single institution and literature review

**DOI:** 10.1101/2020.09.09.20188938

**Authors:** Artem Belyaev, Dmitry Usachev, Marina Ryzhova, Gleb Gulida, Vasilisa Skvortsova, Igor Pronin, Grigory Kobiakov

## Abstract

Adult cerebellar anaplastic astrocytomas (cAA) are rare entities and their clinical and genetic appearances are still ill defined. Previously, malignant gliomas of the cerebellum were combined and reviewed together (cAA and cerebellar glioblastomas (cGB), that could have possibly affected overall results. We present characteristics of 15 adult patients with cAA and compared them to a series of 45 patients with a supratentorial AA (sAA). The mean age at cAA diagnosis was 39.3 years (range 19-72). A history of neurofibromatosis type I was noted in 1 patient (6.7%). An IDH-1 mutation was identified in 6/15 cases and a methylated MGMT promoter in 5/15 cases. Patients in study and control groups were matched in age, sex and IDH-1 mutation status. Patients in a study group tended to have a more frequent multifocal presentation at diagnosis (13% vs. 4.4%) and were associated with longer overall survival (50 vs. 36.5 months), but the difference did not reach statistical significance. In both cAA and supratentorial AA groups presence of the IDH-1 mutation remains a positive predictor for the prolonged survival. The present study suggests that adult cAA constitute a group of gliomas with relatively high rate of IDH-1 mutations and prognosis similar to supratentorial AA. The present study is the first to systematically compare cAA and supratentorial AA with respect to their genetic characteristics and suggests that both groups shows a similar survival prognosis.

## Introduction

Anaplastic astrocytomas of the cerebellum (cAA) are extremely rare entities with just a few case reports or case series published so far [1,3,4,7,9,10,12,14,18,22–25,30,31]. Given the relative weight of the cerebellum (approximately 10% of the brain), one could expect 10% of anaplastic astrocytomas to arise from cerebellum [16]. However, no incidence of cAA has been reported due to its’ utmost scarcity. This is not surprising that literature dealing with malignant gliomas of the cerebellum mostly consists of cerebellar glioblastomas, including both pediatric and adult patients, and brainstem tumours involving cerebellum [5,8,11,15,17,21,32,34].

Meanwhile, cerebellar glioblastomas themselves represent a rare form of neoplasms with reported incidence rate of 0.24-4.1% from all primary glioblastomas [19]. Sporadic articles devoted to the cAA were published decades ago and combined cGB and cAA in the analysis due to insufficient number of patients in each group. In total only 45 cases of cAAs have been documented in the literature (see Table 2). Importantly, previous studies lack any genetic characteristics of cAA and no standard therapeutic approach has been developed or advocated. Here we leverage on the unique opportunity to follow up 15 patients with cAA who received treatment at Burdenko Neurosurgery Center.

**Table 2.** Cerebellar anaplastic astrocytoma patients (global experience)

| ## | Author | Year of publication | Age | Sex | Treatment | OS, wks |
| --- | --- | --- | --- | --- | --- | --- |
| 1 | Budka [3] | 1975 | 41 | F | STR | 1 |
| 2 | Preissig [20] | 1979 | 52 | M | STR, BCNU | 13 |
| 3 | Salazar [23] | 1981 | 8 | F | STR, PFI | 48 |
| 4 |  |  | 6 | F | STR, WBI | 52 |
| 5 |  |  | 7 | M | STR, PFI | 45 |
| 6 |  |  | 10 | M | STR, PFI | 87 |
| 7 |  |  | 16 | M | STR, SCI | 216 |
| 8 |  |  | 13 | M | STR, SCI | 117 |
| 9 |  |  | 10 | M | 2xSTR, SCI | 156 |
| 10 | Alpers [1] | 1982 | 26 | F | STR,RT,BCNU | 1 |
| 11 | Kopelson [15] | 1982 | 15 | M | STR,RT | 459 |
| 12 |  |  | 60 | M | RT | 52 |
| 13 |  |  | 30 | M | STR | 4 |
| 14 |  |  | 67 | M | biopsy | 4 |
| 15 | Zito [34] | 1983 | 77 | F | STR,RT | 12 |
| 16 |  |  | 85 | M | STR | 1 |
| 17 | Steinberg [25] | 1985 | 15 | M | STR,PFI,BCNU,VCR | 30 |
| 18 |  |  | 6 | F | GTR,PFI | 182 |
| 19 | Itoh [9] | 1988 | 1 | F | STR | 1 |
| 20 | Jaskolski [10] | 1988 | 52 | F | 2xSTR, BCNU | 30 |
| 21 | Shinoda [24] | 1989 | 12 | F | STR,RT | 26 |
| 22 |  |  | 12 | F | biopsy | 13 |
| 23 | Chamberlain [4] | 1990 | 14 | ? | 2xSTR, PFI,ChT | 100 |
| 24 |  |  | 21 | ? | 2xSTR, BCNU | 546 |
| 25 |  |  | 44 | ? | STR,PFI,ChT | 221 |
| 26 |  |  | 39 | ? | STR,PFI,ChT | 928 |
| 27 |  |  | 25 | ? | 2xSTR, RT,ChT | 139 |
| 28 |  |  | 33 | ? | STR,PFI,ChT | 26 |
| 29 |  |  | 38 | ? | STR,PFI,ChT | 22 |
| 30 |  |  | 29 | ? | 2xSTR, PFI,ChT | 169 |
| 31 |  |  | 33 | ? | 2xSTR, PFI | 1568 |
| 32 | Marchese [17] | 1990 | 11 | ? | STR, WBI | 115 |
| 33 | Tsunoda [30] | 1992 | 44 | M | STR,WBI,PFI,ChT | 64 |
| 34 | Djalilan [7] | 1996 | 63 | F | STR, PFI | 12 |
| 35 | Walid [31] | 2008 | 14 | M | STR,ChT | >25 |
| 36 | Morgan [18] | 2016 | 47 | M | 2xSTR, RT,ChT | >92 |
| 37 | Kozhuki [14] | 2018 | 75 | M | STR,RT,ChT | >105 |
| 38-45 | Rizk [22] | 1994 | 8 patients |  |  |  |

**Abbreviations:** STR – subtotal resection, GTR – gross total resection, PFI – posterior fossa irradiation, WBI – whole brain irradiation, RT – radiotherapy, ChT – chemotherapy; BCNU – carmustine, VCR – vincristine.

The aim of the present study was to describe genetic and clinical characteristics of 15 adult cAA patients and to compare them both qualitatively and quantitively to those of a series of patients with supratentorial anaplastic astrocytomas with an emphasis to overall survival, multifocal location and IDH-1 mutational status.

## Materials and methods

The database of Burdenko Neurosurgery Center was screened from 2000 to March 2020 and a total of 15 cAA patients met our criteria: 1. AA was located in cerebellar vermis or hemispheres (patients presenting with tumours involving both brainstem and cerebellum were excluded from the study); 2. all patients were evaluated by MRI before admission and underwent maximal safe resection; 3. all patients received adjuvant (chemo- and radiotherapy) in the post-operative period. Biopsy results were reviewed by 3 independent experienced neuropathomorphologists. A consecutive series of 45 adult patients with supratentorial hemispheric anaplastic astrocytomas not involving basal ganglia and/or midline structures were extracted from our database and used as a comparative group. Two patients in the main group presented with multiple lesions (parietal lobe in one case with unconfirmed supratentorial tumour biology and frontal lobe + corpus callosum in another case with glioblastoma histology); in the control group there were also 2 patients with multiple supratentorial AAs. All specimens – both in study and control group – were available for molecular analysis. The groups matched by patient gender, age and IDH-1 mutation status.

## Statistical methods

Survival curves analyses and Cox regression models were performed using R-package (version 3.3.3 (R Core Team, 2017) with package “survival” [27,28].

For the Cox regression investigating the status of the IDH-1 across two groups we performed a regression with clustering patients within each group for the purposes of a robust variance estimation) [29].

For MGMT Cox regression model one of the patients was excluded because of the missing MGMT status leaving 14 patients for the analysis.

## Results

Characteristics of 15 patients with cerebellar anaplastic astrocytomas are presented in Table 3.

**Table 3.** Patients’ characteristics.

| Patients | Personal History | Sex | Age | Tumor Location | Supratentorial Tumor | Surgery | IDH-1 Mutaion | MGMT methylation | Adjuvant treatment | OS (mounth) |
| --- | --- | --- | --- | --- | --- | --- | --- | --- | --- | --- |
| 1 | No | M | 25-30 | H, V | left frontal lobe +CC | GTR,S | + | - | C,R | 9 |
| 2 | No | M | 37-43 | H, V | No | B,S | - | - | C | 48,9 |
| 3 | No | F | 29-33 | H, V | No | GTR | - | - | R | 5,4 |
| 4 | No | F | 45-50 | H | No | GTR | + | + | C,R | 110,6 alive |
| 5 | No | F | 52-56 | H, V | No | STR,S | + | + | C,R | 6,3 |
| 6 | No | M | 40-45 | V | No | GTR | - | - | R | 8,1 |
| 7 | No | M | 70-75 | 1-H<br>2-H,V,BrSt | No | 1-GTR 2-STR | - | - | No | 20 |
| 8 | No | M | 19-23 | V | No | GTR,S | + | - | C,R | 100,4 alive |
| 9 | No | F | 56-60 | H | No | GTR | - | + | C,R | 7,4 |
| 10 | Cr uteri | F | 49-53 | H, V | No | GTR | - | + | C,R | 62,7 |
| 11 | No | F | 38-42 | H | right parietal lobe | GTR | + | - | C,R | 9,3 |
| 12 | No | M | 19-23 | V | No | STR | + | ? | C,R | 159 alive |
| 13 | No | M | 17-21 | H | No | GTR | - | - | C | 30,8 |
| 14 | No | M | 44-48 | V | No | GTR | - | + | C,R | ?? |
| 15 | NF-1 | F | 20-24 | H, V | No | STR | - | - | C,R | 121,8 alive |

**Abbreviations:** NF-1 – neurofibromatosis type I; H - cerebellar hemisphere; V - cerebellar vermis; BrSt – brainstem; CC – corpus callosum; GTR – gross total resection; STR – subtotal resection; B – tumour biopsy; S - shunt placement; C – chemotherapy; R – radiotherapy; OS – overall survival.

## Clinical characteristics

The mean age at diagnosis was 39 years (range 19-72 years). There were 8 men and 7 women. A personal history of neurofibromatosis type I (NF1) was noted in 1 patient (6.7%), as well as a personal history of previous cancer (cervical cancer). Clinical manifestation consisted mostly of intracranial hypertension syndromes (nausea, vomiting) 60% of cases and cerebellar symptoms (gait and writing disturbances) 47% of cases. One of two patients with concomitant supratentorial tumors experienced seizures.

## Histological and molecular characteristics

Biopsy samples were reviewed by 3 independent neuropathomorphologists and patents were included into study group only upon consensus regarding tumour type (in accordance with WHO 2016 classification of CNS tumours). Median Ki67 labeling index was 15% (range 8-50%). Molecular analysis revealed that 6 patients had IDH-1 mutation; methylated MGMT-promoter was identified in 5 patients and H3F3AK27 mutation was revealed in 1 patient.

## Treatment and outcome

One patient underwent tumour biopsy, in 3 patients subtotal resection (STR) was performed, and in 11 patients surgery consisted of gross-total resection (GTR) based upon surgeon’s impression. In one patient repeated surgery was performed in 12 months’ time due to tumour recurrence (no adjuvant treatment was conducted in-between). Notably, the first surgery consisted of GTR, however during the second one only STR was achieved because of brainstem involvement. A CSF-shunt was implanted in 2 patients before admission to our clinic and in one shortly after surgery. All but one patient received adjuvant treatment after surgery that consisted of combined chemoradiotherapy (n=10), chemotherapy alone (n=2) and radiotherapy alone (n=2). Median overall survival (OS) for 14 patients was 50 months with 4 patients surviving for more than 100 months, and being alive at the time of the preparation of this report. One patient dropped out of the follow-up because of moving to another country.

## Comparison with supratentorial anaplastic astrocytomas

Characteristics of patients with cerebellar and supratentorial anaplastic astrocytomas are summarized in Table 2. Patients in the control group were matched in gender, age and IDH-1 mutation status with patients from the study group. Surprisingly, despite virtually ideal match, cerebellar AA had better outcome, than their supratentorial counterparts (OS: 50 months +/− 49.7 vs. 36.5 +/− 35.5 months). However, analysis of Kaplan-Meyer survival curves showed no difference in survival rates (log-rank two-tailed test p = 0.31).

To test for the effects of IDH-1 mutation on the survival outcomes we ran a Cox regression model with group clustering (see Methods) including IDH-1 status, gender, and age as covariates.

Age ad IDH-1 status came out significant predictors. While age has a negative effect on survival (increasing age by 1 year raises the hazard rate by 10.4% (p<0.001) while presence of the positive IDH-1 mutation decreases the hazard rate by 32.1% (p<0.001).

**Table 1.**

|  | coef. | exp(coef) | se(coef) | robust se | z | p |
| --- | --- | --- | --- | --- | --- | --- |
| Age | 0.0426 | 1.0435 | 0.0139 | 0.0124 | 3.42 | 0.00062 |
| IDH1 | -0.3864 | 0.6795 | 0.4001 | 0.0907 | -4.26 | 0.0001 |
| Gender | -0.3082 | 0.7347 | 0.3857 | 0.2975 | -1.04 | 0.30021 |

(the full output for the model)

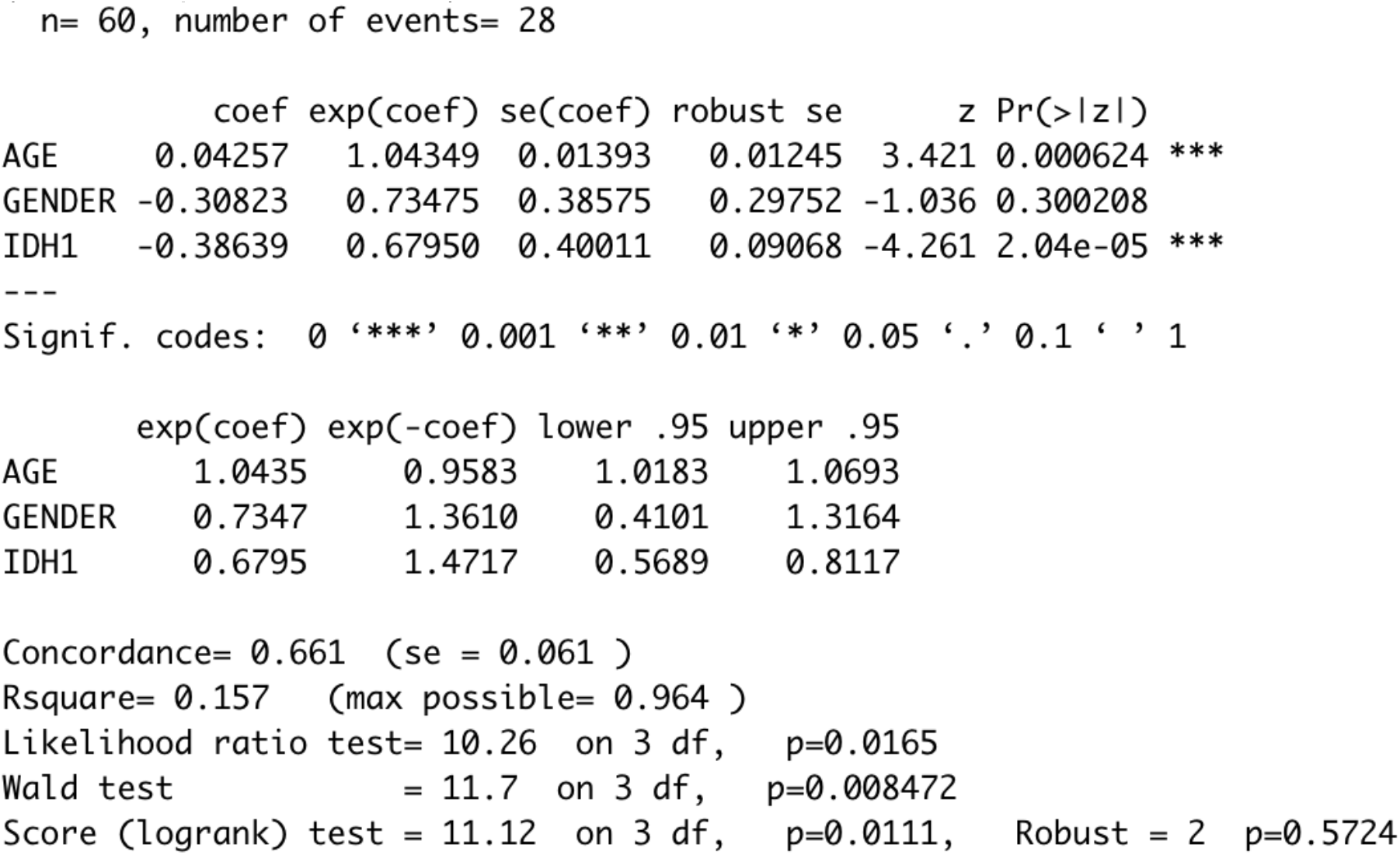

We also looked at the effects of the MGMT-status in cAA group of patients only (since MGMT data were not available in the control group). MGMT-status had a marginal (p = 0.0916) significant negative effect on hazard rates when accounting for gender, age and IDH-1 status but more data are needed to conclude whether positive MGMT mutation might improve survival outcomes.

## Discussion

A thorough literature review revealed just a few articles where “cerebellar anaplastic astrocytoma” term was mentioned, most of them published between 1990 to1998. Because of its’ very limited number, we found it essential to considerate each of them.

Chamberlain et al. [4], described 18 “poorly differentiated gliomas of the cerebellum”, but the sample included a series of glioblastomas (28%), anaplastic astrocytomas (50%) and “mixed gliomas” (22%). Six patients were younger than 18 years old at diagnosis. Median survival rate for 9 cerebellar anaplastic astrocytomas (including 1 pediatric case) was 44 months. Interestingly, 5 out of 18 patients demonstrated tumour relapse: 3 leptomeningeal and 2 parenchymal extracerebellar (parietal lobe and upper cervical cord, respectively). Unfortunately, no information is given regarding the histological type of tumour with extracerebellar relapse.

Rizk et al. [22], also presented a mixed series of cerebellar glioblastomas (n=2) and cerebellar anaplastic astrocytomas (n=8). This study was conducted in late 1970s - early 1990s and based on CT scan tumour appearances. Seventy percent of tumours developed within cerebellar vermis, 30% in hemispheres. Authors did not state median OS for either cGB or cAA, but 4 out of 8 cerebellar anaplastic astrocytomas were still alive by the time of article submitting without any signs of recurrence with a median follow-up of 7 years.

Paper of the most interest published by Djalilian et al. [7] presented an analytic review of malignant gliomas of the cerebellum and was based on 78 cases (37 cAA and 41 cGB), although only 7 of them were of authors’ clinical experience. Median survival for patients with cAA was 32 months. Obvious conclusion is made that patients undergoing surgical resection and receiving radiation therapy demonstrated better survival rate. Despite the fact that the article combined both pediatric and adult cases, it provided a systematic review of malignant astrocytomas of the cerebellum revealing in particular 19 adult cAA patients (OS in this group was 53.8 months).

Since all those papers were published decades ago, no information regarding genetic profile of malignant cerebellar gliomas was reported.

In this study we presented a series of 15 consecutive adult patients with cerebellar anaplastic astrocytomas treated in a single institution for over 20-year period. Survival analyses was performed with respect to patients age and IDH-1 mutational status, as well as MGMT methylation profile. We used control group of supratentorial AA as a “perfect match” with no difference in age, sex and IDH-1 mutation status.

Our results are aligned with previous studies demonstrating relatively prolonged survival in a group of cAA patients comparing to their supratentorial counterparts: in our group of cAA patients’ OS was 50 months vs. 36.5 months in control group of supratentorial AA patients, but this difference did not reach statistical significance due to high variability in our sample. Median OS in a review published by Djalilian for 19 adult cAA patients was very similar – 32 months. [7] OS in our control group was similar to the data published before: Strowd et al. [26] demonstrated OS for AA in the temozolomide era as 36,7 months. For this comparison, it is noteworthy that both patients harboring additional supratentorial tumour (in case 1 AA, case 11 of unknown histology) had very short overall survival – 9 and 9.3 months, respectively. Probably, not only multiple location itself, but also a different tumour behavior could affect OS in those patients. Interestingly, the eldest patient in the group demonstrated 20-months OS despite the absence of any adjuvant treatment; he underwent repeated surgery after 12 months, but then deteriorated due to brainstem involvement by the tumour.

Rizk et al. [22], reported that 4 out of 8 cAA patients presented with more than 7 years of followup without sings of recurrent disease.

This higher survival rate of patients with cAA when comparing to supratentorial AA remains a puzzling phenomenon: malignant cerebellar tumors hypothetically should be associated with a heavier burden due to their fast expansion and building-up mass-effect in a small compartment of posterior fossa, containing critical life-supporting structures of the brainstem. Moreover, tumour may involve brainstem itself with catastrophic consequences. Despite all those well-recognized facts, cAA cases pertain a better prognosis than supratentorial tumours of the same histological type. It is also true for cerebellar glioblastomas: Yang et al. [33] presented a series of 28 cGB patients with OS 14.3 months. In a series of 202 cGB collected from SEER database and presented by Babu et al. [2] OS both in cGB group and in supratentorial GB (sGB) group was similar (7 months). Multicenter study of cerebellar GBM conducted by Weber et al. [32] and based upon 45 cGB cases also suggested that prognosis of infratentorial GB was not different from supratentorial tumours with median OS of approximately 10 months. However, in all abovementioned series cGB groups were younger than sGB and that could have possibly affected OS rates. In our group age factor was carefully controlled by strict matching. No clear clarification of this phenomenon has been proposed so far.

One of the promising explanations – as well as for the rarity of malignant gliomas in the cerebellar - may be a substance P deficit in the cerebellar tissue. This amino acid peptide neurotransmitter is active throughout the cerebrum and brainstem, but not in the cerebellum. Substance P signaling has been shown to play a contributory role in glioblastoma development. [11]

Quite an unexpected result of the present study is a very high incidence of IDH-1 mutations in the study cohort (40%). In a similar study with 17 cerebellar GB conducted by Picart et al. [19] none of the reported tumours demonstrated IDH-1 positive status. It is a well-known fact that for supracerebellar gliomas IDH-1 mutation pertains better prognosis [6,13]. Overall, our results confirm this tendency for the whole cohort (study + control groups). Unfortunately, we failed to demonstrate it separately for cAA patients due to the group small sample size and it will need further investigation.

Our results also pointed towards a potentially promising role of methylated MGMT promoter status in influencing prolonged survival in cAA patients, but the effect remains marginal and will require future replications.

Finally, patients in the study group had more frequent multifocal presentation (13% vs. 4.4% in the control group). One could argue that cerebellar affection is only a sign of the late stage of disease. To align with this assumption, both patients in the study group with multifocal gliomas demonstrated OS much lower than average (9 and 9.3 months, respectively, compared to 50 months).

Despite its limited sample-size and the absence of large-scale molecular analysis, the present study suggests that cAA do not correspond to a homogeneous entity but constitute a heterogeneous subgroup of anaplastic gliomas. We believe further accumulation of data for cAA with subsequent meta-analysis might shed light on cAA characteristics and possibly refine classification of these tumors.

**Figure 1.**
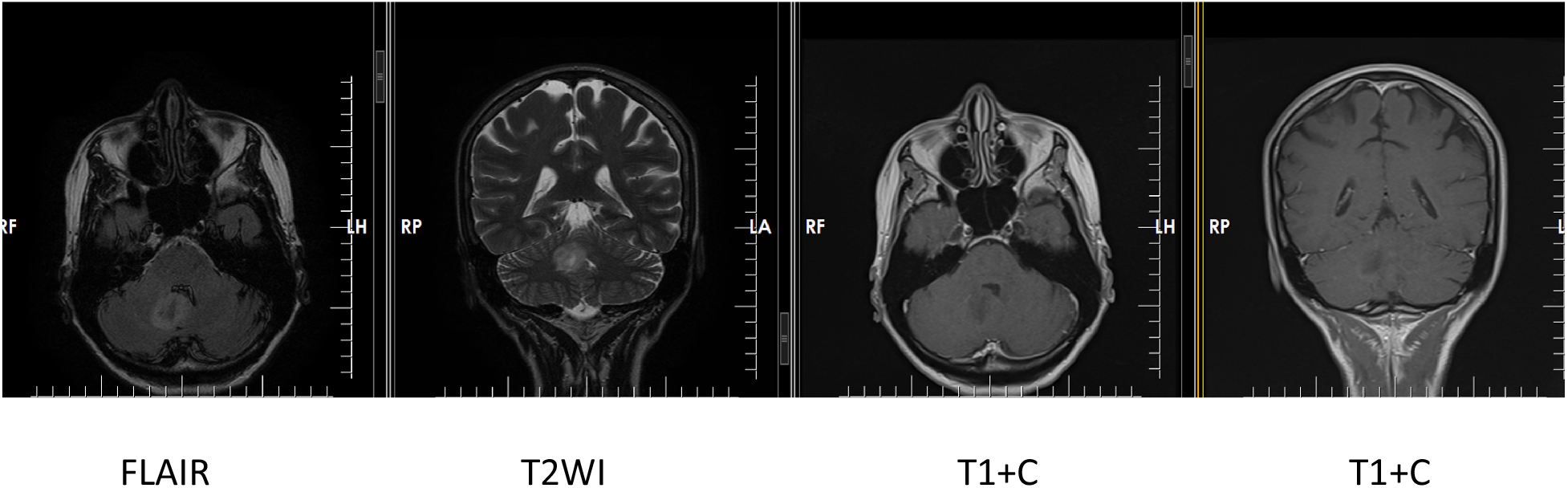
Patient 45-49 years, pre-op MRI.

## Data Availability

All data is available online through medical journal websites and PubMed search engine

https://thejns.org

https://pubmed.ncbi.nlm.nih.gov

https://www.springer.com/journal/701

https://www.springer.com/journal/11060

https://www.sciencedirect.com/journal/world-neurosurgery

